# Clinician’s opinions, views and expectations on radiological reports: A multicentre cross-sectional study in Dar es Salaam region

**DOI:** 10.1101/2025.11.23.25340851

**Authors:** Amon Danda, Zuhura Nkrumbih, Lulu Fundikira, Jerry Hella

## Abstract

**Background:** Radiology reports are the primary communication tool between radiologists and clinicians. They guide diagnosis, treatment, and follow-up care, yet in many settings there are no formal feedback mechanisms to improve their quality. In Tanzania, where medical imaging capacity is rapidly expanding, evidence on clinicians’ perceptions of report quality remains scarce.

**Methods:** A multicenter cross-sectional study was conducted in the Dar es Salaam region involving 320 clinicians. Data were collected using a self-administered questionnaire and Google Forms. Descriptive statistics were applied to summarize clinicians’ responses, and chi-square tests were used to assess associations.

**Results:** Most clinicians reported positive opinions regarding the understandability (79%), clarity (92%), and relevance (91%) of radiology reports. A standardized report format was preferred by 83% of respondents. Preferences for the location of positive findings were nearly evenly split (54% favored reporting them first vs. 46% at the respective anatomical location). Use of standardized lexicons or grading systems was supported by 93% of clinicians, while 55% valued clarification of radiological terms in the conclusion. Overall, 87% were satisfied with report content and 86% expressed general satisfaction, recognizing radiology reports as critical for clinical decision-making. Report delays and inadequate timely communication of urgent findings were identified as key shortcomings.

**Conclusions:** Clinicians in Dar es Salaam expressed overall satisfaction with the clarity, relevance, and structure of radiology reports, while highlighting the need for timely delivery and improved direct communication of critical findings.

## Introduction

Radiology reports are the primary mode of communication between radiologists and clinicians, serving an essential role in diagnosis, treatment planning, and follow-up care. These reports are also integral parts of patient records and may carry medicolegal implications [1]. High-quality reports are expected to be concise, clinically relevant, clearly structured, and timely. Report writing is regarded as the cornerstone of radiological practice [2], with training and experience being central to the generation of quality reports [1,3]. Conversely, poorly structured or inaccurate reports may compromise patient safety, contribute to mismanagement, and expose healthcare providers to medicolegal risks. They may also result in unnecessary interventions, wasted time, and increased healthcare costs for both patients and insurance providers [4].

Globally, studies assessing clinicians’ perspectives on radiology reporting have been conducted in Asia, Europe, and the Americas. However, evidence from Africa and Australia remains sparse, with only one study reported from Kenya within East and Central Africa [5]. In Tanzania, substantial investments in imaging services have expanded access to advanced technologies such as Computed Tomography (CT) scanners, accompanied by the training of radiology personnel. Over the past decade, the country has witnessed significant growth in both diagnostic equipment and skilled professionals [6]. Despite these advancements, little is known about clinicians’ perceptions of the quality of radiology reports. Radiology reporting remains a predominantly one-way communication tool, and the absence of structured feedback mechanisms limits opportunities for professional development and continuous improvement [7].

This study aims to address this knowledge gap by assessing clinicians’ perspectives and expectations regarding radiology reporting in Dar es Salaam, Tanzania. Findings will provide context-specific evidence to inform strategies for improving report quality, enhance collaboration between radiologists and clinicians, and contribute to the regional literature on radiology reporting practices in resource-limited settings.

## Methods

### Study design

A multicenter cross-sectional study was conducted in both public and private hospitals in Dar es Salaam, providing a snapshot of clinicians’ opinions, views, and expectations regarding radiology reports during the study period.

### Study sites

The Dar es Salaam region comprises five districts (Kinondoni, Ilala, Temeke, Ubungo, and Kigamboni) with a population of 5,383,728 residents. According to the 2022 census by the National Bureau of Statistics, the region has 76 hospitals and health centers. Of these, 24 facilities have at least one radiologist and a CT scan machine, as recorded in the Ministry of Health facility register. Ilala hosts the largest number (11), followed by Kinondoni (10), Ubungo (2), Temeke (1), and none in Kigamboni. Among the 24 facilities, nine are publicly owned. These hospitals were stratified into public and private categories, and six were randomly selected from each group.

**Table 1:** Randomly selected study sites.

| **Public owned hospitals** | **Private owned hospitals** |
| --- | --- |
| i. Muhimbili National Hospital Upanga | i. CCBRT Hospital |
| ii. Muhimbili Orthopedics Institute (MOI) | ii. Ranbinsia Memorial Hospital |
| iii. Muhimbili National Hospital Mloganzila | iii. Bochi Hospital Limited |
| iv. Temeke Regional Referral Hospital | iv. Hurbert Kairuki Memorial Hospital |
| v. Mwananyamala Regional Referral Hospital | v. Agha Khan Hospital Dar es Salaam |
| vi. Amana Regional Referral Hospital | vi. Sali international Hospital |

### Study population

The study population included all specialists working in the selected hospitals in the Dar es Salaam region.

### Study duration

Recruitment of study participants was conducted between 1^st^ June 2024 to 31^st^ March 2025.

**Figure 1.**
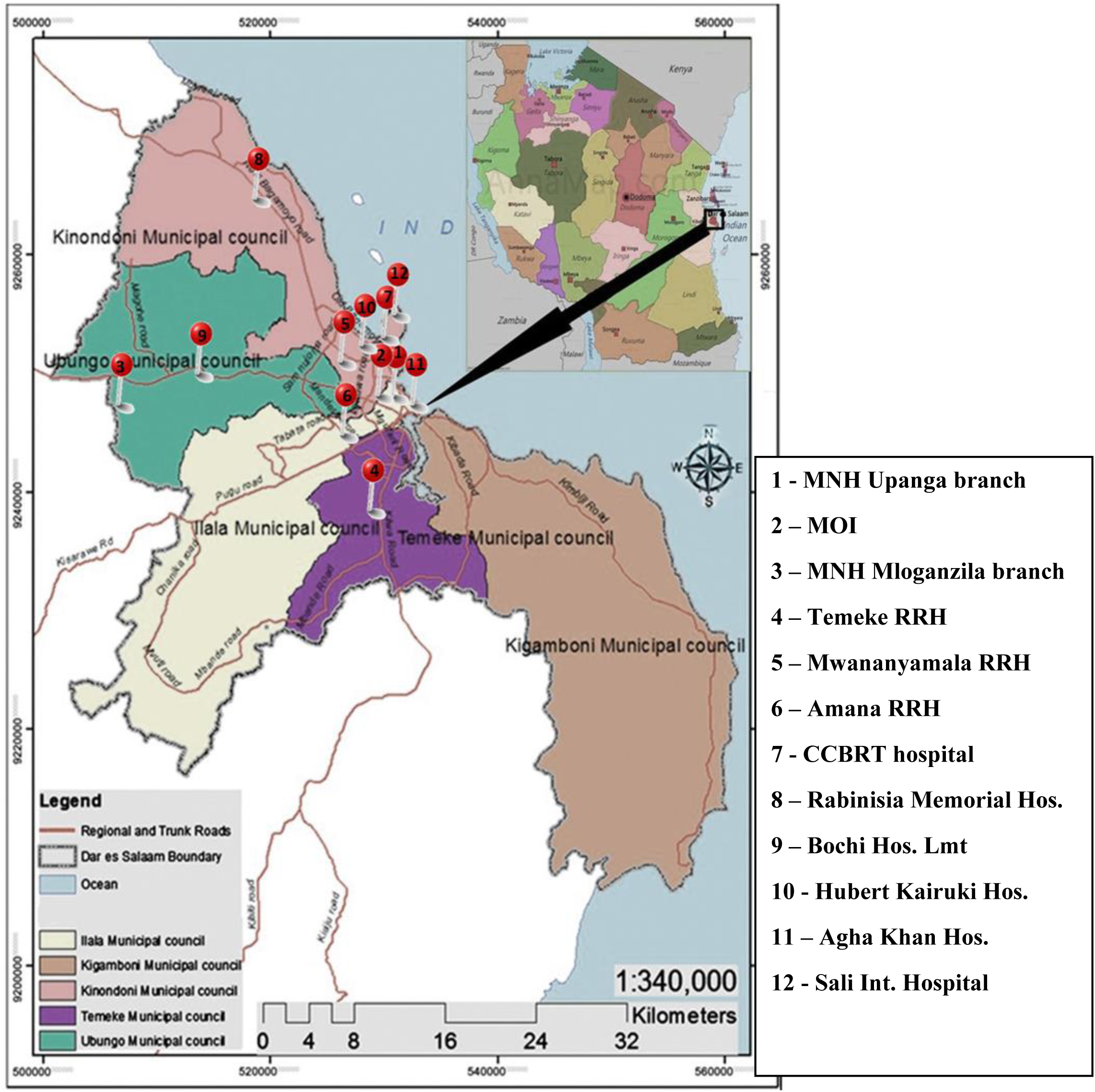
Map of Dar es Salaam city with randomly selected study sites.

### Sample size consideration

The sample size was determined using the Fisher equation for sample size calculation.

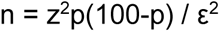

Where:

N= Desired sample size
z= standard normal deviate=1.96 for 95% confidence level
p = expected proportion with characteristic of interest.
ε = margin of error (precision)

A recent paper conducted in Kenyatta Hospital in Kenya in 2020 showed the following characteristics of interest; Relevance and valuability of radiology reports 77.8%, preference for a structured report – 76.6% and 29.3% received verbal communication on emergence cases.

Thus, the expected proportion of characteristic of interest (P)

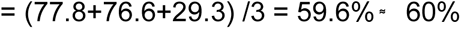

= (77.8+76.6+29.3) /3 = 59.6% ͌ 60%

Margin of error (ε) is set at 5%

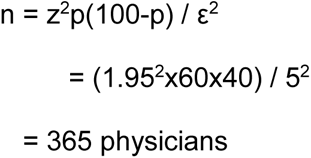

### Inclusion and exclusion criteria

The following were the inclusion and exclusion criteria.

Inclusion criteria

i. Clinicians attending patients for at least the past 6 months.
ii. Clinicians receiving at least 3 reports per week

Exclusion criteria

i. Clinicians working in radiology or Interventional radiology department.

There were excluded due to conflict of interest as their work was being assessed.

### Data collection procedure/sampling technique

The questionnaire was largely adapted from the COVER studies conducted in Europe, America, and Asia (4,11,14,19), with adjustments made to reflect the Tanzanian context and culture. A pilot study involving 5% of the sample size was conducted to assess the validity and reliability of the adapted tool, ensuring that the information collected was relevant and accurate. Minor wording adjustments were made accordingly.

Twelve research assistants were trained on the study protocol, including procedures for obtaining consent, ensuring data quality, and conducting follow-up. Data were collected using a self-administered questionnaire, either in the presence of a research assistant or at the convenience of the participant. In addition, a Google electronic form was created to capture responses from clinicians who were not physically present at the hospital but were willing to participate. All eligible clinicians working in the randomly selected hospitals in the Dar es Salaam region were invited to take part in the study.

### Research instruments

The questionnaire comprised of four parts, Section A contained demographic data, Section B assessed clarity and relevance, Section C assessed clinician’s views on style and content and Section D assessed clinician’s expectations and need.

A five-point Likert scale was used to grade responses. A Likert scale comprises a range of options that allowed clinicians to express their opinions, views and expectations. It was used to gauge the degree of clarity, frequency, relevance, acceptance and need. A five-point Likert scale had two positive and negative options of varying degrees of agreement/disagreement and a neutral one. There were also free text sections to capture opinions/views/expectations limited by Likert scale.

### Data management and analysis

Data was entered and analysed using statistical software Epi Info^TM^ version 7.2.0.1 for windows. Descriptive statistics were used to summarize demographic characteristics of the study participants. Frequencies and percentages were used to present categorical variables.

Clinicians’ opinions on clarity and relevance were tethered to a five-point Likert scale as such frequency tallies were obtained, interpreted and analyzed accordingly. Views on reporting style and content were assessed using multiple-choice questions thus frequency, proportions and mean were used in analysis. Lastly, expectations and need were assessed using both multiple-choice and five-point Likert scale questions as such frequency tallies in relation to predictor variables, proportions and means were used in interpretation and analysis. Statistical test Chi square was used to determine whether there was association between predictor variables and expectations of clinicians on radiology reports. The last two sections had open-ended questions that captured recommendations and expectations outside the scope of the questionnaire. Responses were categorized accordingly and analyzed.

### Definition of variables

Predictor variables are radiology report, area of medical specialization, number of years of medical practice, sex, Hospital ownership and number of reports received per week.

Outcome variable are clinician’s Likert scores and responses.

### Ethical issues

The research proposal was approved by the Muhimbili University of Health and Allied Sciences (MUHAS) Institutional Review Board (IRB) (MUHAS-REC-05-2024-2208), and the approval was forwarded to the respective hospitals for internal authorization.

Research assistants provided prospective participants with detailed explanations about the study and the implications of participation. The study posed no potential risks, as it did not involve any invasive or therapeutic procedures. Informed consent was obtained from all participants and documented on the questionnaire cover page. No personally identifiable data were collected, ensuring the protection of participants’ privacy and confidentiality.

## RESULTS

The study involved 328 clinicians (90% of the estimated sample size). We excluded 8 participants due to incompletely filled questionnaires.Thus, 320 clinicians were included in the study.

**Table 2:** Characteristics of the study population.

|  | Frequency<br>(n) | Percent<br>(%) |
| --- | --- | --- |
| **<u>Sex</u>** |  |  |
| - Male | 197 | 62 |
| - Female | 123 | 38 |
|  | **320** | **100** |
| **<u>Education background</u>** |  |  |
| - Specialists and fellows | 262 | 82 |
| - Superspecialists | 58 | 18 |
|  | **320** | **100** |
| **<u>Number of years of practicing</u>** |  |  |
| - 1-10 years | 204 | 64 |
| - 11 and above | 116 | 36 |
|  | **320** | **100** |
| **<u>Hospital ownership</u>** |  |  |
| - Public | 216 | 68 |
| - Private | 104 | 32 |
|  | **320** | **100** |
| **<u>Departments</u>** |  |  |
| - Emergency | 16 | 5 |
| - Internal medicine | 80 | 25 |
| - Surgery | 92 | 29 |
| - Obstetrics and gynecology | 59 | 18 |
| - Pediatrics | 39 | 12 |
| - Others | 34 | 11 |
|  | **320** | **100** |

- Median of number of radiology reports read per week by clinicians – 10 (IQR 3-20)
- Average number of years of medical practice – 10.4 years with SD 7.9, with a range of 1-46 years.

### Specific objectives

#### i) Clinician’s opinions on clarity and relevance of radiology reports

**Figure 2:**
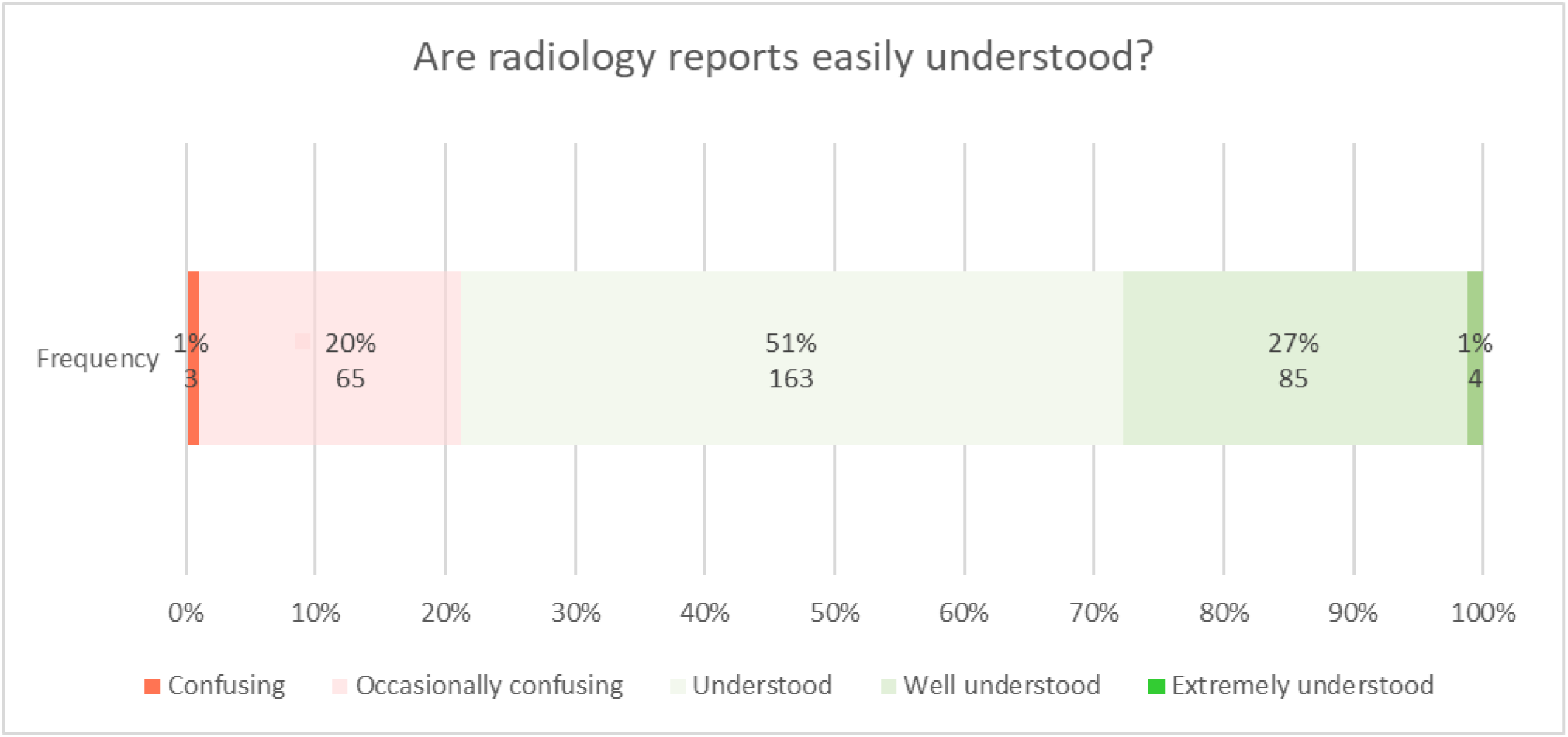
Clinicians’ understanding of radiological reports.

Majority of the clinicians 252(78.75%) reported they understood radiology reports.

**Figure 3:**
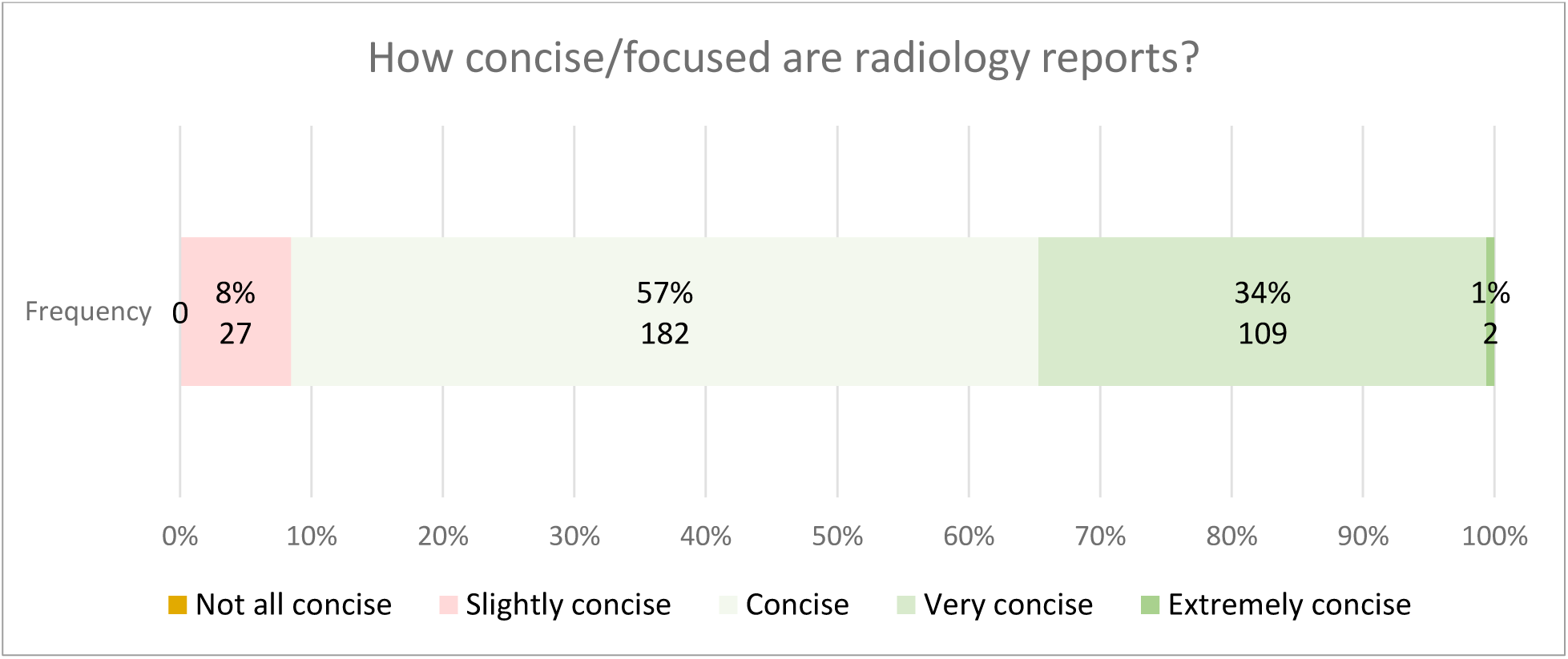
Conciseness of radiology reports among clinicians.

Most of the clinicians, 293 (91.57%) reported radiology reports are concise/focused.

**Figure 4:**
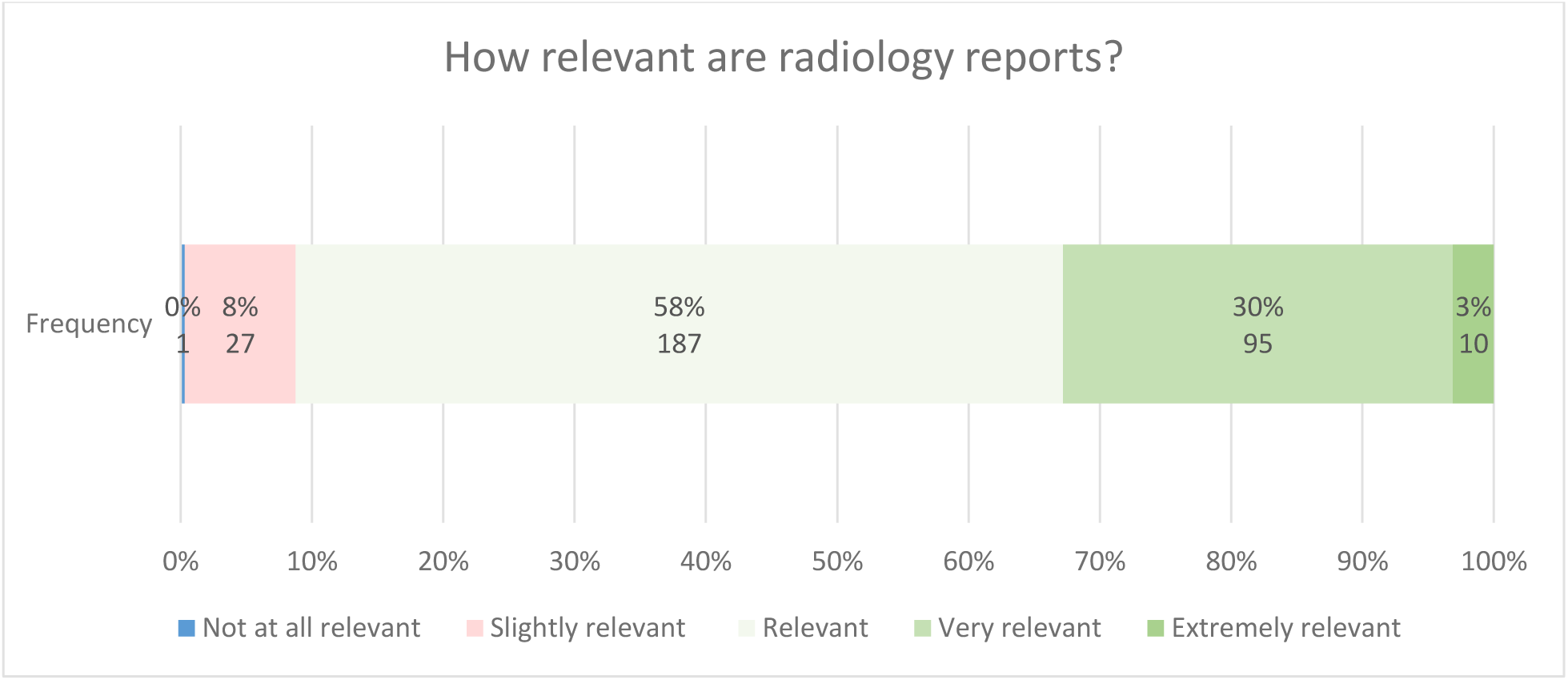
Relevancy of radiology reports are among clinicians.

Most of the clinicians 292 (91.26%) reported radiology reports are relevant.

#### ii) Clinician’s views on reporting style and content of radiology reports

**Table 3:** Preferred format of radiology report among clinicians.

| *Preferred format of radiology report* | *Frequency (n)* | *Percent (%)* |
| --- | --- | --- |
| - *Standard itemized format (3,4)* | 266 | 83.13 |
| - *Prose format (3,4)* | 29 | 9.06 |
| - *No opinion* | 25 | 7.81 |
| **Total** | **320** | **100.0** |

Majority of the clinicians 266(83.1%) prefer standard itemized report with subheading of each organ/system.

**Figure 5:**
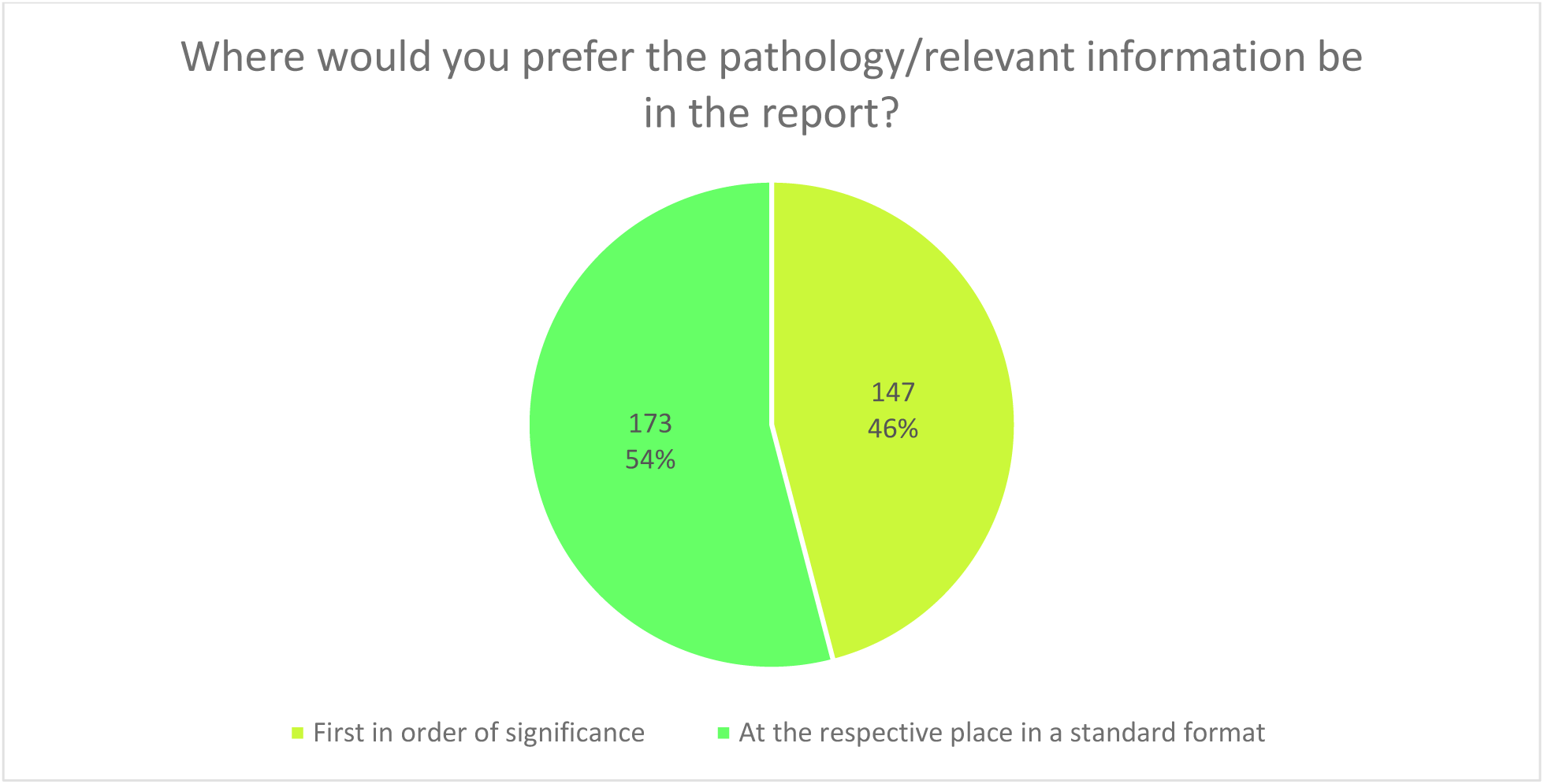
Preferred location of positive finding in radiology report.

Preference for the location of pathology/relevant information is almost equally distributed.

**Figure 6:**
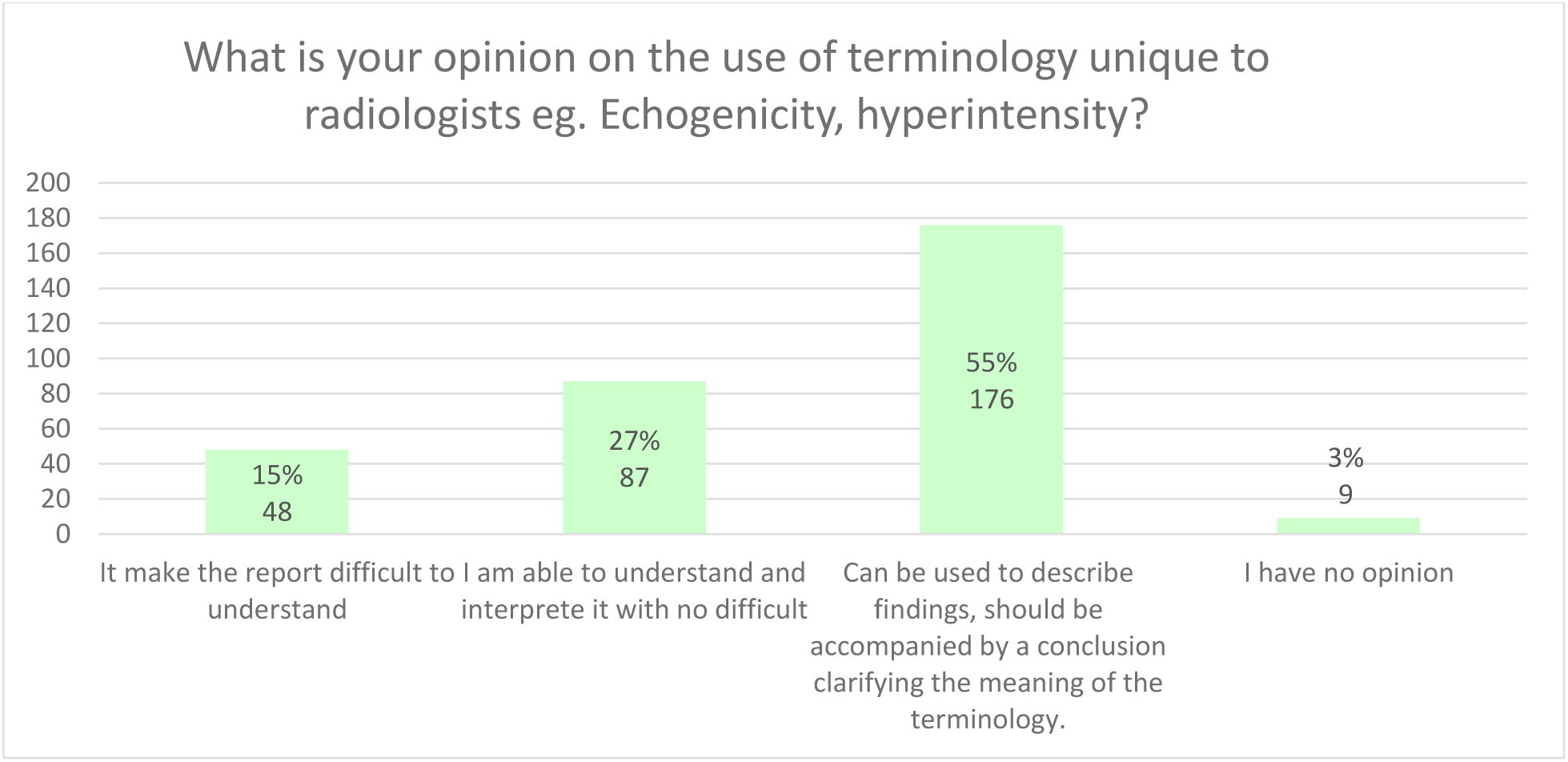
Clinicians’ opinions on the use of terminologies unique to radiology.

Most clinicians, 176(55%) prefer radiological terms to be used in the description, accompanied by a conclusion clarifying the meaning.

**Figure 7:**
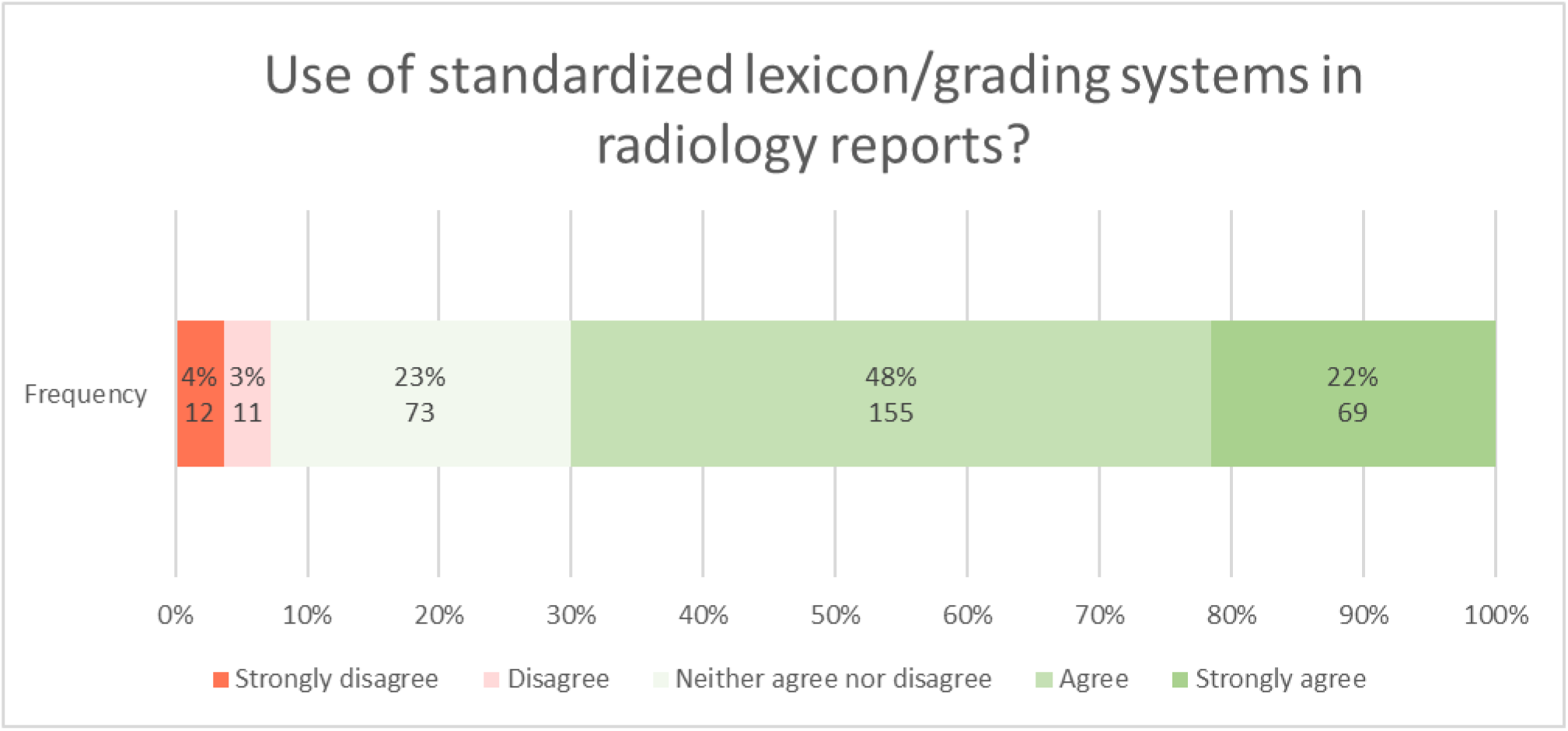
Clinicians’ preference on the use of lexicons/grading systems.

Most clinicians 224(70%) prefer the use of standardized lexicon/grading systems in radiology reports.

**Figure 8:**
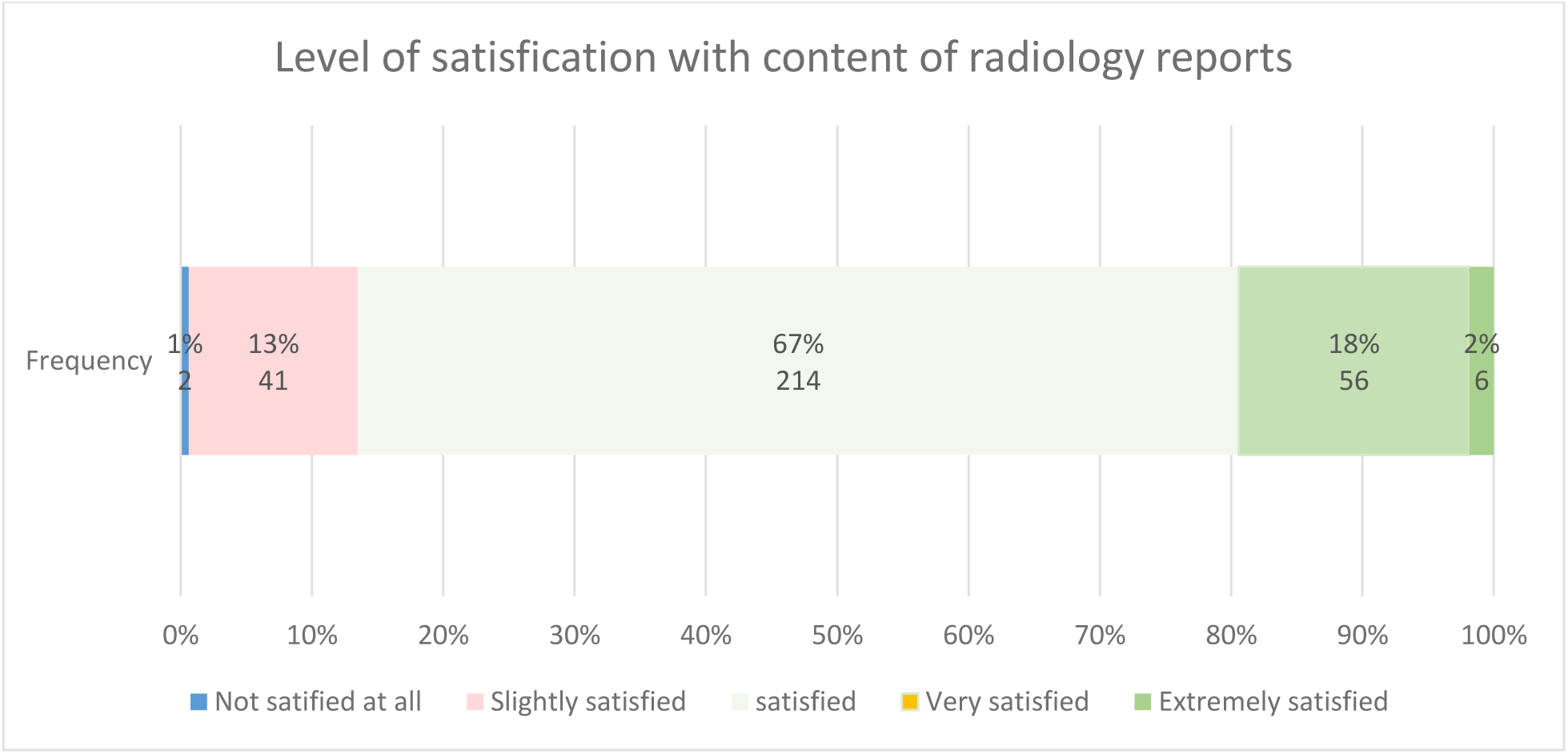
Clinicians’ satisfaction with content of radiology reports.

270(86.81%) of clinicians are satisfied with content of radiology reports.

**Figure 9:**
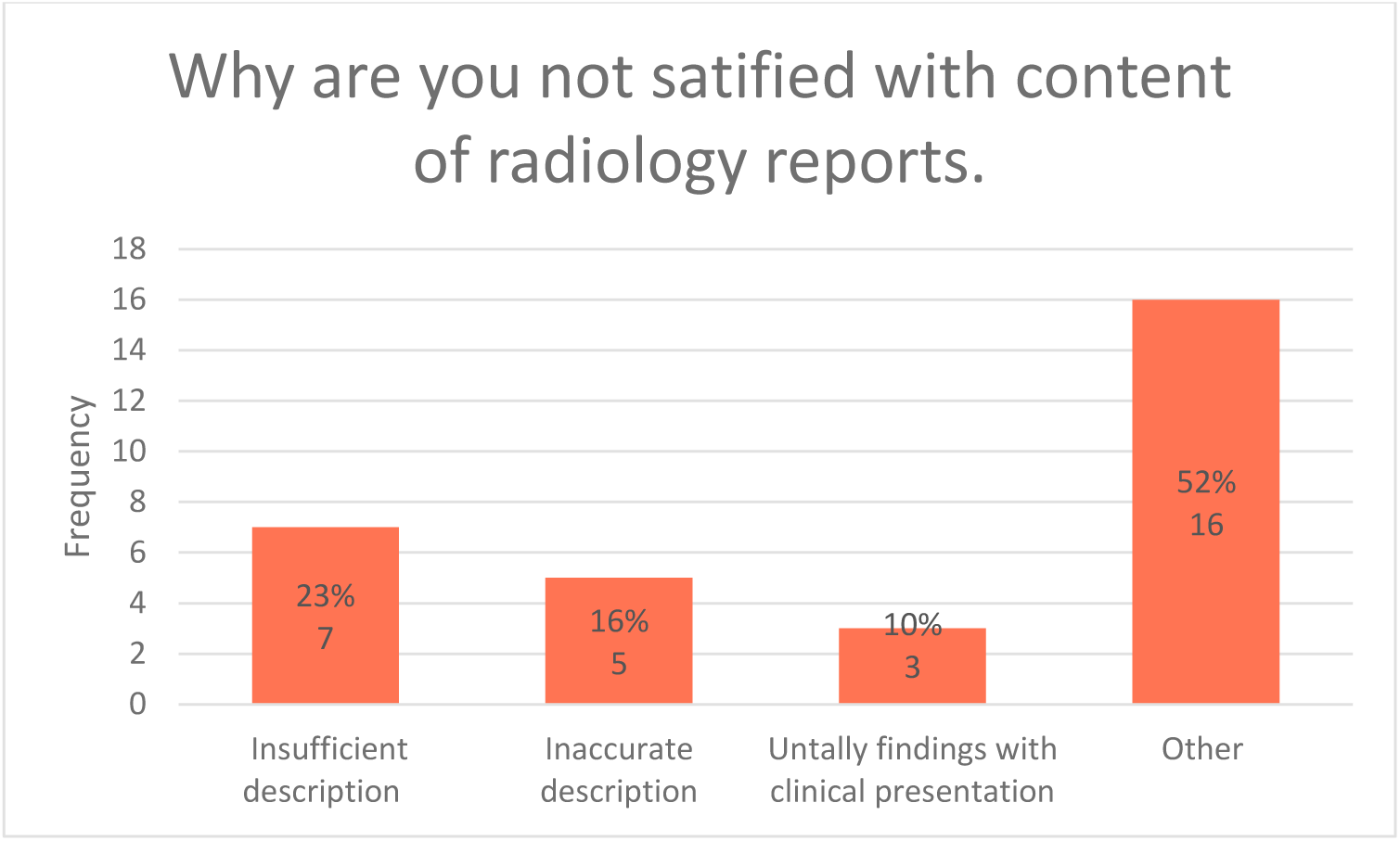
Reasons for clinicians’ dissatisfaction with content of radiology reports.

#### iii) Assessment on whether radiological reports met clinicians’ expectations and need

**Figure 10:**
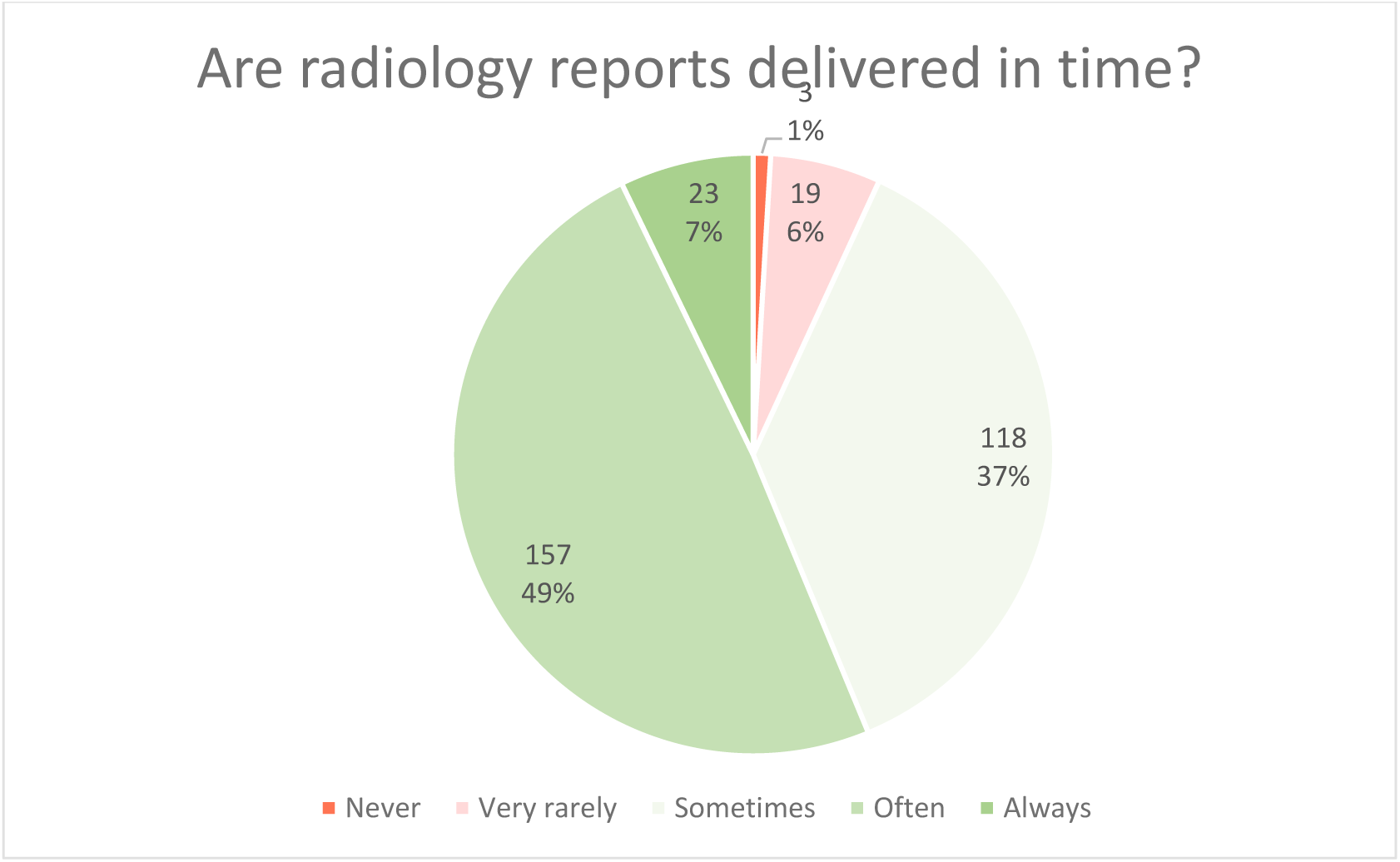
Timely delivery of radiology reports.

22 (7%) of the radiology reports are delivered late as reported by clinicians and 180 (56%) are often/always on time.

**Figure 11:**
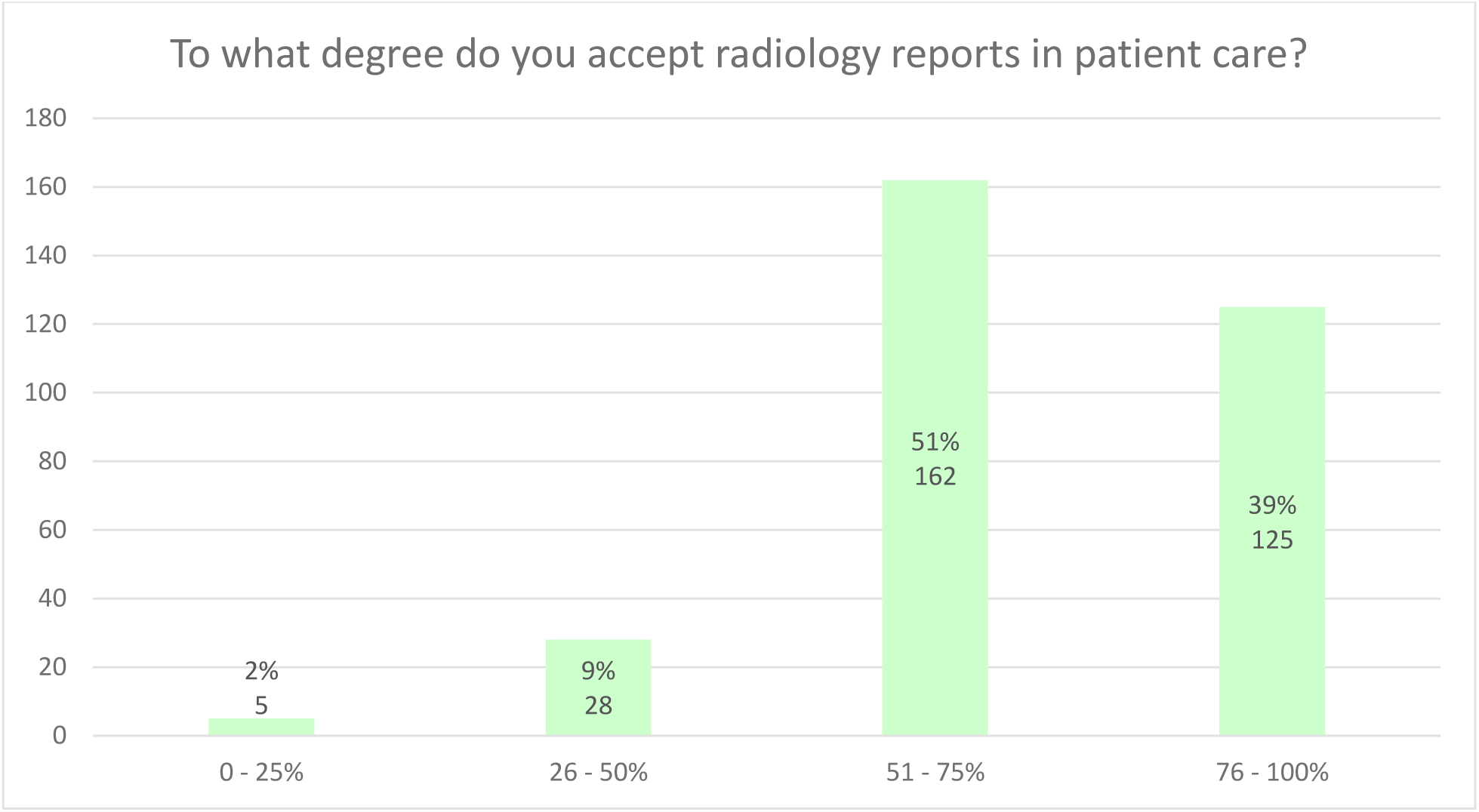
Acceptance of radiology reports in making medical decisions.

287(90%) of clinicians accept and use radiology reports 51-100% of the times, in making medical decisions.

**Table 4:** Clinicians’ expectations of radiology reports.

| Do radiology reports meet your expectations? | Frequency (n) | Percent (%) |
| --- | --- | --- |
| - Positive | 274 | 85.6 |
| - Negative | 21 | 6.6 |
| - Undecided | 25 | 7.8 |
| **Total** | **320** | **100.0** |

Majority of the clinicians 274 (85.63%) had positive expectations with radiology reports.

**Figure 12:**
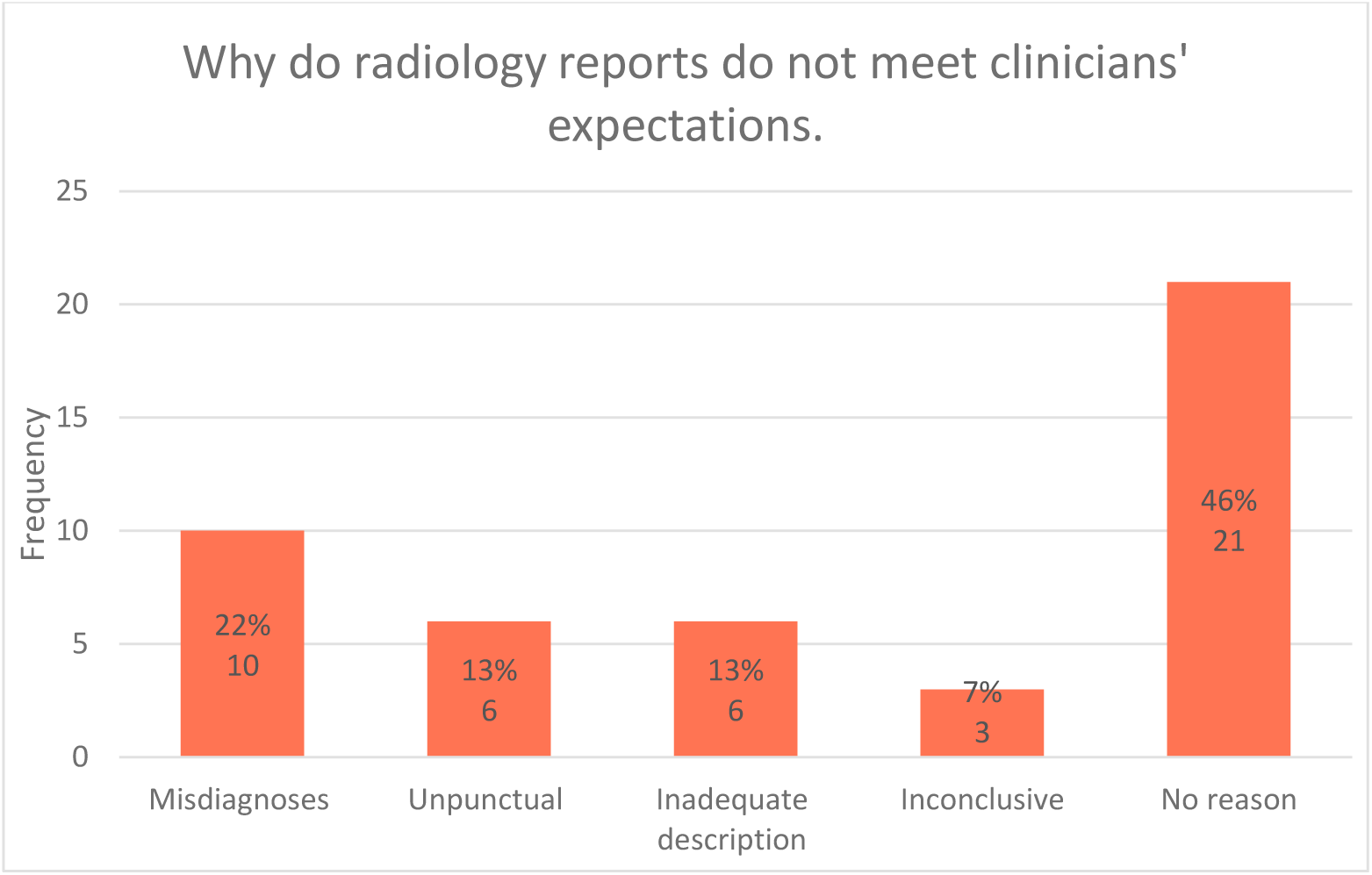
Reasons for clinicians’ disappointment with radiology reports.

Out of the 46 disappointed/undecided clinicians, 21 (46%) did not have a reason.

**Table 5:**
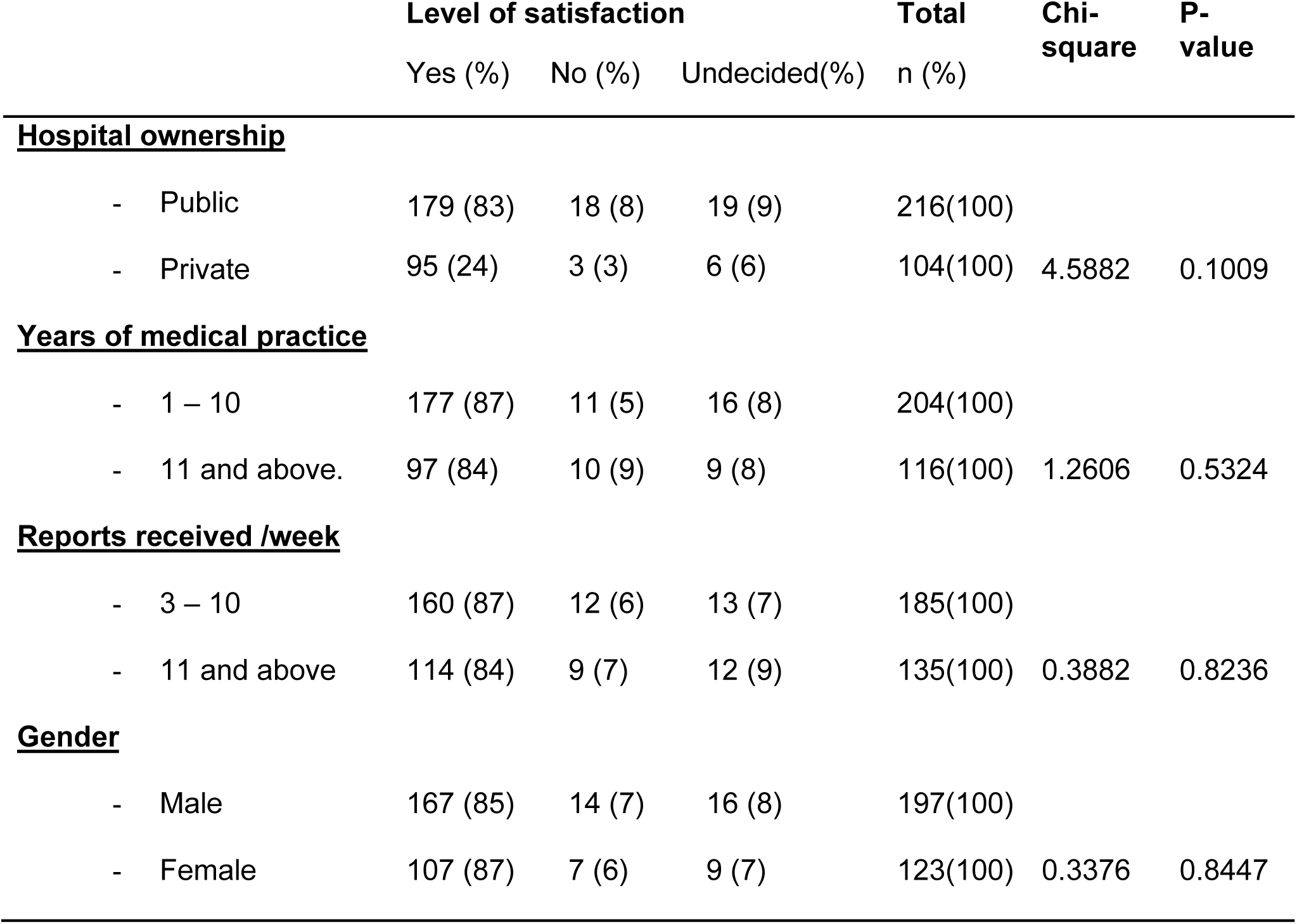
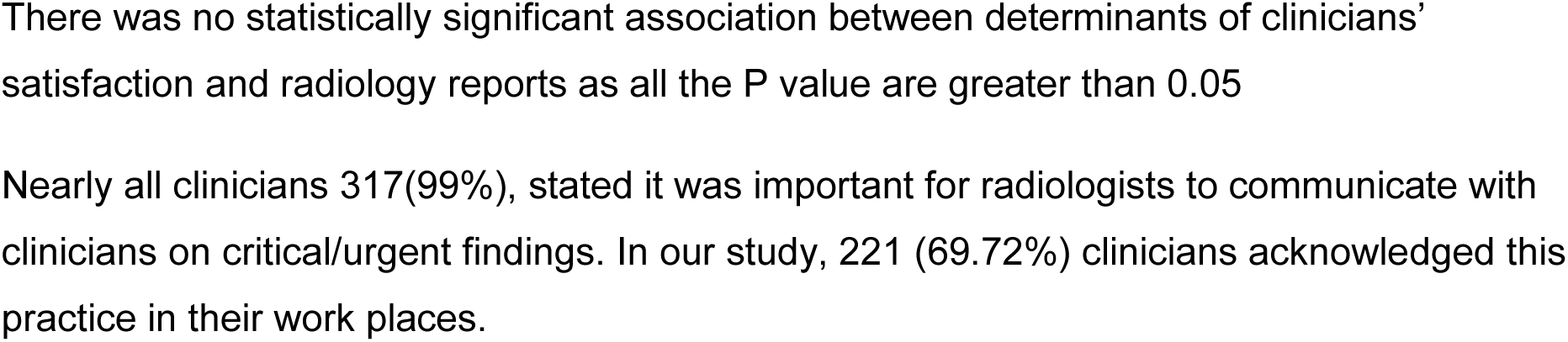
Associated determinants of level of satisfaction with radiology reports.

There was no statistically significant association between determinants of clinicians’ satisfaction and radiology reports as all the P value are greater than 0.05

Nearly all clinicians 317(99%), stated it was important for radiologists to communicate with clinicians on critical/urgent findings. In our study, 221 (69.72%) clinicians acknowledged this practice in their work places.

## Discussion

This study reaffirmed the critical role of radiology reports in patient care, consistent with previous research conducted across Africa, Asia, Europe, and the Americas (4,11,15,18,19). The majority of clinicians’ opinions, views, and expectations in the Dar es Salaam region aligned with findings from other parts of the world, suggesting that the ongoing national expansion of medical imaging corresponds with perceived report quality and value. Notably, the study also identified unexpected patterns of association and highlighted shortcomings in timely report delivery and communication of urgent findings, as per ACR standards. Clinicians’ opinions on report clarity and relevance were evaluated under three themes: understandability, conciseness, and relevance. Understandability findings were comparable to the Royal Blackburn Hospital study in the United Kingdom (12) and the binational cross-sectional study in the Netherlands and Flanders (4), likely attributable to the multicenter design and large sample sizes. In contrast, studies from single-center, high-volume urban hospitals, such as Santa Tomas Hospital in the Philippines and Civil Hospital Karachi, reported lower levels of clarity (2,11). Relevance was relatively high in our study compared with Kenyatta National Hospital in Kenya and Civil Hospital Karachi (2,14), potentially due to our multicenter design encompassing both public and private secondary and tertiary hospitals, unlike the single-center public tertiary studies. This aspect has not been widely assessed in studies from Asia, Europe, or the Americas.

Regarding report style and content, international consensus supports standardized reporting per ACR guidelines (1). Clinicians in our study preferred a standardized format at a similar rate to the Netherlands and Flanders study (4), both large multicenter studies across public and private hospitals. The Kenyatta National Hospital study reported slightly lower preference (14). Only the Kenyatta study assessed clinicians’ preferred placement of positive findings, with results ambivalent between first or respective positions (14), likely reflecting specialty-specific interests in the report.

Most clinicians in our study emphasized that radiology-specific terminology should be accompanied by clear conclusions; this was observed at higher rates in Civil, Santa Tomas, and Kenyatta studies, possibly due to tertiary hospital workload pressures (2,11,14,16).

Preference for standardized lexicons or grading systems exceeded 70% in our study and the Santa Tomas study but was lower in the Kenyatta study, likely reflecting differences in clinician composition, with specialists and fellows predominating in the former studies and general practitioners in the latter (11,14).

Overall, the majority of clinicians’ expectations were met, with 86% expressing satisfaction with radiology reports as a key component in medical decision-making. These results align with findings from Santa Tomas, Kenyatta, rural South Dakota, Massachusetts General Hospital, and the Netherlands/Flanders studies (4,15,18). These findings cut across number of study centers, study sample sizes, study location and population pressures. It likely attributable to improved report quality that answers clinicians’ daily medical concerns and inter-specialty trust built in the process.

Slightly lower dissatisfaction rates in our study compared with the Kenyatta and Santa Tomas studies may be due to the former being a multicenter design that included of both public and private hospitals, which captured a broader range of clinical needs (11,14,15,16). Generally, special needs from a given medical specialty not captured in the standard radiology report may also have a contribution.(15,16).

Timely report delivery was suboptimal in our study (56%), Civil Hospital study (48%) and Kermanshah University of Medical Sciences study (38%) unlike in the rural sparsely populated midwestern South Dakota (2, 18, 21), likely reflecting workflow constraints in densely populated urban centers. However, these observations did not take into account the nature of investigation and the turnaround time in the respective centers.

Conversely, communication of urgent or critical findings was relatively high (69.7%), similar to the small sample sized (70 clinicians) single center Federal University of Triangulo, Uberaba Hospital study in Brazil (7), whereas Kenyatta and Civil Hospital studies reported lower rates, possibly due to being single-center wit high patient loads (2,14).

No significant association was observed between clinicians’ satisfaction determinants and radiology reports, contrasting with the Belfast NHS Trust study, which reported a positive correlation between clinician seniority and report value (22). This discrepancy may relate to the fact that 64% of clinicians in our study were junior practitioners with fewer than 10 years of experience.

Key strengths of this study include its multicenter design, inclusion of both public and private hospitals at secondary and tertiary levels, and recruitment of a representative sample of clinicians across all participating centers, enhancing the generalizability of the findings

## Limitations

The study experienced a low response rate, and the targeted sample size was not fully achieved, introducing a potential risk of non-response bias, as the views of responders may not fully represent those of non-responders. However, given the relative homogeneity of the clinician group in terms of knowledge, training, and professional culture, the impact of non-response bias is likely reduced compared to studies involving the general population.

As a quantitative study, the data collected were limited, preventing in-depth exploration of underlying issues. Additionally, the study assessed radiology reports in general, rather than specific imaging modalities, which limits the applicability of the findings to modality-specific opinions and expectations, these highlight an opportunity for future research.

## Conclusion

This study reaffirmed the pivotal role of medical imaging in patient care. Findings indicate that the majority of clinicians in Dar es Salaam region hold positive opinions regarding clarity (understandability and conciseness) and relevance of radiology reports.

Clinicians exhibited nearly equal preferences regarding the positioning of positive findings, while most favored the use of a standardized, itemized report format, use of standardized lexicons or grading systems where appropriate, and clarification of radiological terminology in the conclusions.

Overall, clinicians’ expectations and needs were largely met, with most expressing satisfaction and acknowledging radiology reports as a key pillar in medical decision-making. Nonetheless, a minority reported dissatisfaction with the timeliness of report delivery and communication of critical or urgent finding

## Data Availability

The data that support the findings of this study are available from the author upon request.

## Acknowledgements

The authors acknowledge the Department of Radiology and Imaging of MUHAS for enabling the study to be conducted. I appreciate the invaluable support I received from Dr Zuwena Kassim (MNH Mloganzila), Dr Hanee Mohammed (Sali international hospital), Dr Mpemba Gerald (MNH Upanga), Dr Vaileth Kalinga (MOI), Dr Bernadetha Nguvila (Amana RRH), Dr Festo Shayo (CCBRT), Dr Meekson Mambo (Bochi Hosp. Ltd), and all the colleagues and collaborators who assisted in the data collection process.

